# May we overcome the current serious limitations for distributing reconstituted mRNA vaccines?

**DOI:** 10.1101/2021.03.09.21253129

**Authors:** Santiago Grau, Olivia Ferrández, Elena Martín-García, Rafael Maldonado

## Abstract

There is an urgent need to ameliorate the transport of the reconstituted vaccines to the vaccination sites to improve the COVID-19 vaccination campaigns. The maintenance of the integrity of the mRNA of the different COVID-19 reconstituted vaccines after continuous movement at room temperature during at least three hours ensures the safety of a ground transportation.

## Maintext

On December 27, 2020, the vaccination campaign against COVID-19 began in Spain with the Pfizer-BioNTech vaccine. On January 12, 2021, this vaccination campaign was enlarged in Spain by the introduction of a second vaccine commercialized by Moderna (1). Both vaccines are composed of mRNA and the main difference lies in the lipid nanoparticles that integrate the vaccines with the purpose of protecting the mRNA integrity and facilitate its entry inside the cells. Due to this different lipid content, the Pfizer-BioNTech vaccine requires special storage in freezers between -90°C to -60°C, whereas the Moderna vaccine should be stored between -25°C to -15°C (2). Pfizer-BioNTech vaccine reception in hospitals entails their reconstitution and re-storage at temperatures between 2°C and 8°C with an expiration of 5 days, while the expiration is only 2 h at room temperature. Moderna vaccine can be stored refrigerated between 2°C to 8°C for 30 days and between 8°C to 25 °C for up to 12 h before use. When the first dose is withdrawn from the vial, both vaccines should be held between 2°C to 25°C and must be discarded 6 h later.

The regional Departments of Health are responsible in Spain for centralizing vaccine storage for its subsequent distribution to leading hospital centers, which act as distributors for their reference area. The initial COVID-19 vaccination campaign promoted in Spain consists in a first stage covering the nursing homes population, and later health personnel stratified according to their risk. The second stage has included people according to their degree of vulnerability, followed by personnel belonging to essential critical services. This campaign requires a rapid and efficient distribution of the vaccines from the hospitals to the different vaccination centers. Vaccination teams were recruited, mainly nursing personnel, who got the vaccines from their hospital pharmacy departments, where they are delivered, always keeping the traceability conditions of storage by computer devices (Sirius Storage Monitoring Software). The staff of the nursing homes and the other institutions in charge of vaccination maintains the strict recommendations for handling the vials, ending the process with vaccine administration and the patients’ clinical follow-up. In a scenario of increasing SARS-CoV2 spreading with new highly infective variants and with prospects that predict a dramatic cumulative mortality rate in the next months that could reach 3,600,000 worldwide in the month of June (3), it is mandatory to improve the vaccination rate. Thus, more agile handling of centralized preparation of individualized doses of vaccines, maintaining the best conditions of vaccine preparation, transport, and traceability could improve the vaccination process’s efficiency.

Today, it is recommended that reconstituted vials of the Pfizer-BioNTech and Moderna vaccine or product in syringes should not be transported to avoid unnecessary movement that could alter the integrity of the mRNA (2,4). Reconstituted vaccines should be sent for ‘just in time use’ as part of a planned vaccination clinic. Indeed, once the syringes with the vaccine doses have been prepared, their transport to other places is presently avoided, which represents a major limitation for the rapid spread of vaccination (4). Therefore, there is an urgent need to ameliorate the possibilities to transport the reconstituted vaccines from the centralized preparation centers to the vaccination site. However, no information is currently available about the consequences of movement in the integrity of the reconstituted vaccines. We have now analyzed Pfizer-BioNTech and Moderna vaccine integrity under different movement conditions and provide novel information that may be crucial to improve the vaccines’ distribution to the target population.

The Hospital del Mar (Barcelona, Spain) acted as a reference hospital and the pharmacy department as a vaccine distributor. Doses not administered for expiration or potential loss of microbiological traceability were returned to this pharmacy department. During the first weeks of vaccination, several vials and syringes of Pfizer-BioNTech COVID-19 and Moderna vaccines were returned to the pharmacy department by the vaccination teams after exceeding the expiration time or potential loss of microbiological traceability, such as vials falling to the ground. We selected for this analysis only the vials and syringes with the potential loss of microbiological traceability, discarding those returned due to exceeding the expiration time. Based on these criteria, syringes of reconstituted Pfizer-BioNTech or Moderna COVID-19 vaccines (0.5-2 ml) were prepared in a laminar flow chamber (Hospital del Mar pharmacy department) to be subjected to a stability analysis (Neuropharmacology-Neurophar) to evaluate the impact of movement conditions on mRNA integrity. For this purpose, samples were exposed for 180 min at room temperature (21±1C°, 55±10% of humidity) under different conditions. A first group (n=12 samples from Pfizer-BioNTech and n=17 samples from Moderna COVID-19 vaccines) remained without movement at room temperature for 180 min to mimic the manufacturer’s vaccine conditions use after reconstitution. A second group (n=10 samples from Pfizer-BioNTech and n=16 samples from Moderna COVID-19 vaccines) was exposed to continuous movement during 180 min in swing shaker (Ovan SW-3DE) at 40rpm with an inclination angle (± 8°) to mimic the movement that may occur in ground transportation under the most unfavorable road conditions. Such a period of 180 min could guarantee ground transportation from 180 to 300 km in Europe and North America’s road conditions. This distance would ensure the distribution of the vaccine after reconstitution to the institutions that do not have the appropriate facilities to do it. A third group (n=8 samples from Pfizer-BioNTech and n=14 samples from Moderna COVID-19 vaccines) was exposed to 1 min Vortex (Scientific Industries SI™ Vortex-Genie™ 2) at 3200rpm every 20 min during 180 min, a massive shaking reported to impair RNA integrity (5). We compared the integrity of the samples before and after exposure to these room temperature conditions. Thus, a small amount (2 µL) of the fresh samples (n=12 samples from Pfizer-BioNTech or n=23 samples from Moderna COVID-19 vaccines) that remained without movement during 180 min was extracted before this period under sterile conditions and immediately analyzed.

Microfluidic measurements to analyze mRNA integrity were performed using Agilent 2100 Bioanalyzer (Agilent Technologies, Santa Clara, USA) with the RNA 6000 Nano LabChip kit and the assay Eukaryote total RNA Nano (Genomics Core Facility, University Pompeu Fabra, Spain). Results were generated and analyzed with the Bioanalyzer 2100 Expert Software (Version B.0210.SI764) and manual integration combined with smear analysis was used to define regions following the Bioanalyzer user guide. A negligible mRNA degradation was revealed in the Pfizer-BioNTech and Moderna COVID-19 vaccine samples exposed to room temperature during 180 min with or without swing shaker, similar to the fresh samples before exposing to these conditions. Indeed, the RNA fractions area under the original mRNA peak was similar in these three experimental groups with very low values of fluorescence units (FU) (Figure 1a and 2a) that correspond to less than 5% of original mRNA degradation for both Pfizer-BioNTech and Moderna samples (Figure 1b and 2b). Any possible product of the original mRNA’s degradation should be identified in these fractions of lower molecular weight.

**Figure 1.**
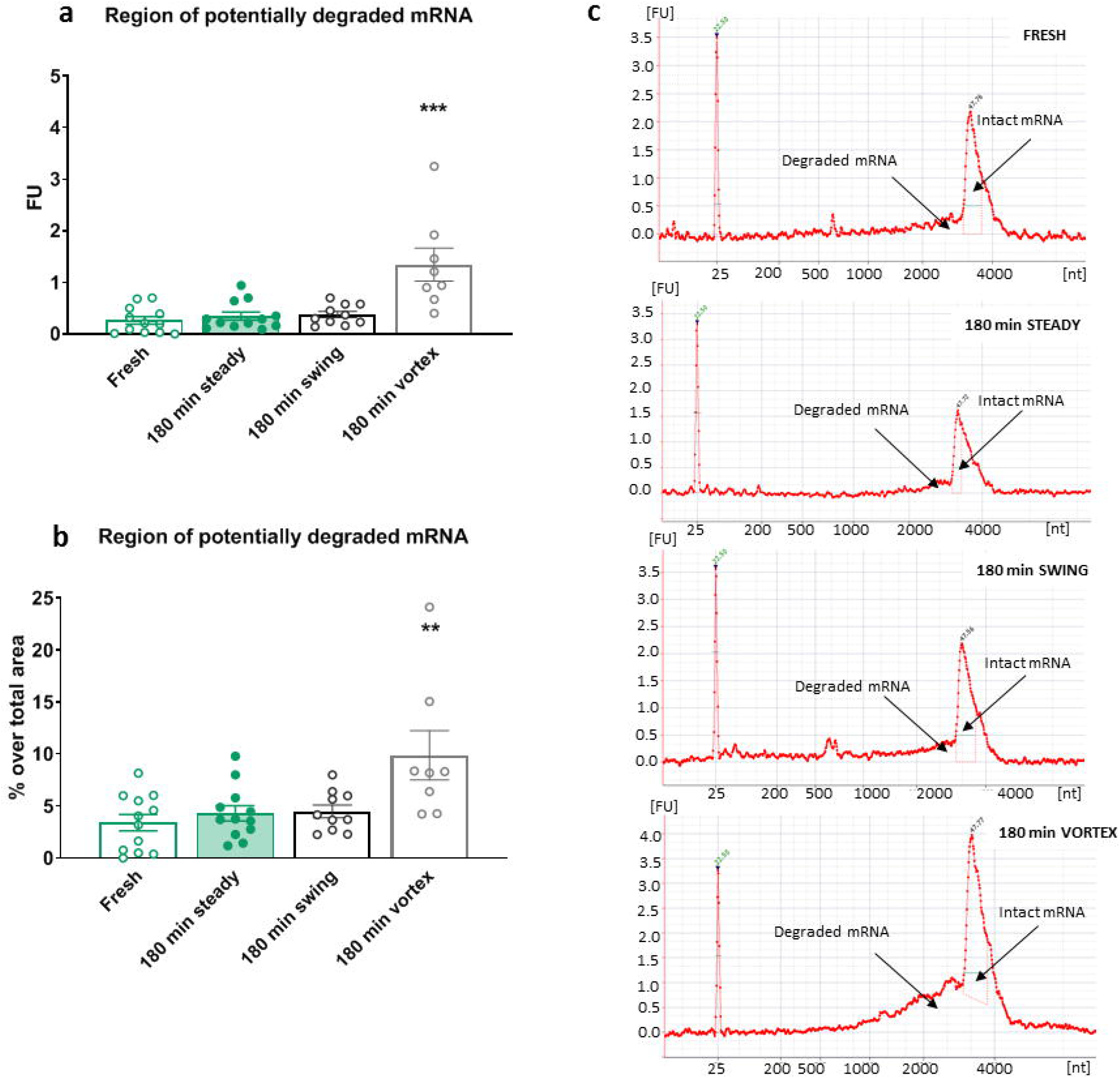
Analysis of mRNA integrity of reconstituted Pftzer-BioNTech COVID 19 vaccines. a) Measurement of the region of poientially degraded mRNA in fluorescence units (FU) of the samples immediately analyzed (fresh), or after 180 min without movement (180 min steady), continuous swing shaker movement (180 min swing), or vortex shaking (180 min vortex). Individual values of FU with the mean ± SEM are represented; one-way ANOVA, F(3,38)=13.39, p<0.00 1, Newman-Keus *(N-K)* post hoc test ***P < 0.001 vortex vs the remaining groups (n=8-12). b) Percentage over the region’s total area of potentially degraded mRNA of the samples immediately analyzed, or after 180 min without movement, continuous swing shaker movement, or vortex shaking. Individual values of FU percentage with the mean ± SEM are represented; one-way ANOVA, F(3,38)=5. 772, p<0.01, *N-K* post hoc test **P < 0.01 vortex vs the remaining groups (n=8-12). c) Representative electropherograms expressed in FU of RNA integrity for different mRNA samples detailing the regions that indicate potentially degraded and intact mRNA peaks in the samples immediately analyzed, or after 180 min without movement, continuous swing shaker movement, or vortex shaking.

**Figure 2.**
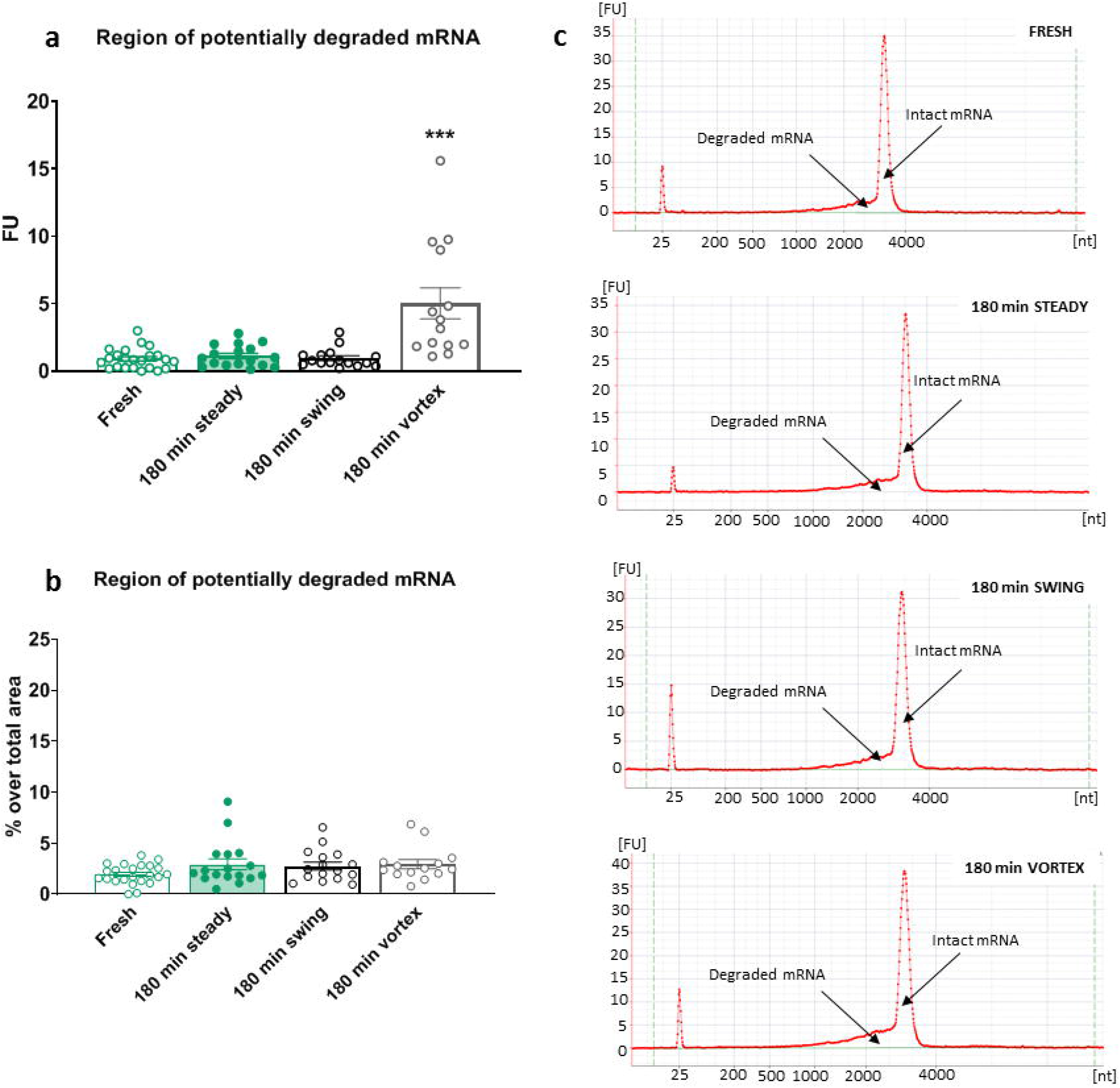
Analysis of mRNA integrity of reconstituted Moderoa COVID 19 vaccines. a) Measurement of the region of potentially degraded mRNA in fluorescence units (FU) of the samples immediately analyzed (fresh), or after 180 min without movement (180 min steady), continuous swing shaker movement (180 min swing), or vortex shaking (180 min vortex). Individual values of FU with the mean ± SEM are represented; one-way ANOV A, F(3,66)=14.77, p<0.001, Newman-Keus *(N-K)* post hoc test ***P < 0.001 vortex vs the remaining groups (n=14-23). b) Percentage over the region’s total area of potentially degraded mRNA of the samples immediately analyzed, or after 180 min without movement, continuous swing shaker movement, or vortex shaking. Individual FU percentage values with the mean ± SEM are represented; one-way ANOVA, F(3,66)= 1. 745, n.s. (n=14-23). c) Representative electropherograms expressed in FU of RNA integrity for different mRNA samples detailing the regions that indicate potentially degraded and intact mRNA peaks in the samples immediately analyzed, or after 180 min without movement, continuous swing shaker movement, or vortex shaking.

In contrast, the integrity of the mRNA samples of Pfizer-BioNTech and Moderna vaccines exposed to repeated Vortex shaking was significantly impaired. Indeed, the area of the RNA fractions under the original mRNA peak was 1.34±0.31 FU (P<0.001) for the Pfizer-BioNTech samples (Figure 1a), which corresponds to 9.87±2.36% of original mRNA degradation (P<0.01) (Figure 1b). In agreement, the area of the RNA fractions under the original mRNA peak of the Moderna samples after Vortex shaking was 5.03±1.16 FU (P<0.001) (Figure 2a), which corresponds to only 2.95±0.45% of original mRNA degradation (NS) (Figure 2b) due to the higher RNA fraction of the peak corresponding to non-degraded mRNA in the Moderna (147.26±17.80 FU) than in the Pfizer-BioNTech (10.92±1.00 FU) samples.

Therefore, a continued moderate movement at room temperature of the reconstituted Pfizer-BioNTech and Moderna vaccines that mimics ground transportation does not impair the mRNA’s quality since this quality was similar without movement and did not differ from the original quality of the fresh samples. As expected, the mRNA contained in these reconstituted vaccines may be degraded under the standard conditions reported to impair mRNA integrity (5), although the degradation after this massive shaking was only moderate mainly in the case of the Moderna vaccine. Therefore, exposure of the Pfizer-BioNTech and Moderna COVID-19 reconstituted vaccines to continuous movement mimicking ground transportation during 180 min at room temperature does not impair mRNA quality. The permanence of the mRNA’s integrity under these experimental conditions suggests that the Pfizer-BioNTech and Moderna COVID-19 reconstituted vaccines and subsequently fractionated in syringes in pharmacy departments could be more easily distributed than initially expected using ground transportation.

It is extremely important that the population gain confidence in the face of vaccination against COVID-19 (6), which may be conditioned by excessively long waiting periods to receive the vaccine. The stability of the Pfizer-BioNTech and Moderna COVID-19 reconstituted vaccines after continuous movement at room temperature may improve the efficiency in the administration of the vaccines, which may lead to shorter and more homogeneous vaccination in cities and rural areas. Indeed, the possibility of preserving the mRNA after medium-distance ground transportation of reconstituted and fractionated syringes of both vaccines may optimize the preparation of final vaccine doses and the distribution efficiency. This could particularly improve the distribution of the vaccines to benefit institutions that currently cannot receive these vaccines due to the lack of facilities for locally reconstitute the samples and that are not located close to the centers available to reconstitute the vaccines. This ground transportation would be crucial for quickly reaching the different points indicated by the health systems to obtain homogeneous vaccination coverage of the population in a short time frame.

## Data Availability

The data that support the findings of this study are openly available.

## Key Messages

- There is an urgent need to ameliorate the possibilities to transport the reconstituted vaccines from the centralized preparation centers to the vaccination sites to improve the COVID-19 vaccination campaigns.
- The integrity of the mRNA of the COVID-19 reconstituted vaccines after continuous movement at room temperature is maintained for at least three hours.
- An appropriate knowledge of this stability may improve the efficiency in the administration of the vaccines, which may lead to shorter and more homogeneous vaccination in cities and rural areas.

